# An emerging trend in STI Management: Antibiotic STI Post-Exposure Prophylaxis Prevalence and Determinants in Recent Surveys

**DOI:** 10.64898/2026.02.06.26345737

**Authors:** Bill Berners-Lee, James Bell, Caisey Pulford, Dana Ogaz, Hamish Mohammed, Ross Harris, John Saunders

**Affiliations:** Blood Safety, Hepatitis, STI & HIV Division, UK Health Security Agency; NIHR HPRU in Blood Borne and Sexually Transmitted Infections; Modelling Division, UK Health Security Agency

## Abstract

**Objectives:** This study aimed to estimate the prevalence of antibiotic post-exposure prophylaxis (STI PEP) use among gay, bisexual and other men who have sex with men (GBMSM) in England, and to identify demographic and behavioural factors associated with STI PEP use among early adopters.

**Methods:** We synthesised individual-level data from nine surveys conducted between 2019 and 2024. To harmonise variables that differed subtly across surveys, we generated composite indicators for STI diagnosis, chemsex, and condomless sex. Multiple imputation was used to address partial missingness; participants missing core sociodemographic data were excluded. The primary outcome was self-reported STI PEP use. We calculated the prevalence of use in each survey and assessed associations using mixed-effects logistic regression. Composite variables were included as random effects if they were found to have an interaction with survey.

**Results:** Across nine surveys with 14,142 participants, STI PEP use prevalence ranged from 3.61% (95% confidence interval (CI): 2.78–4.68%) in RiiSH 2020 to 13.05% (11.85–14.36%) in RiiSH 2024. Use was higher among those aged 40–44 (adjusted odds ratio (aOR) = 1.26; 95% CI: 1.00–1.58) and lower in those under 25 (aOR = 0.56; 0.39–0.82) or over 65 (aOR = 0.64; 0.41–0.99), compared to ages 30–34. Transgender women (aOR = 3.87; 1.75–8.50), Black (aOR = 1.85; 1.34–2.56) and Asian (aOR = 1.43; 1.09–1.88) individuals, and people living with HIV (aOR = 3.40; 2.69–4.31) had higher odds of use, compared with cisgender male, White, and HIV negative individuals, respectively. Chemsex (aOR = 2.15; 1.80–2.56), condomless sex (aOR = 1.59; 1.32–1.92), and recent STI diagnosis (aOR = 1.83; 1.34–2.51) were also associated with use.

**Conclusion:** STI PEP use was more common among those practising behaviours associated with higher STI risk. However, participation and sampling biases limit finding generalisability, particularly to underrepresented groups such as women, ethinic minorities and heterosexuals. This analysis will provide a baseline from which to estimate the impact of the recommendation of doxycycline for STI post-exposure prophylaxis in the UK.

## Introduction

Syphilis is a sexually transmitted infection (STI) caused by the *Treponema pallidum* bacterium (TP). If untreated, syphilis is associated with serious morbidity and can result in congenital syphilis when untreated through pregnancy[1]. There has been an increasing trend in infectious syphilis diagnoses in England since the early 2000s, with the 9,535 diagnoses in 2024 being the highest annual number reported since the 1940s[2]. A persistently high burden of syphilis is something that has been observed globally, as evidenced by data from the Global Burden of Disease Study 2019[3] and various regional studies[4-5].

It is in this context that doxycycline post-exposure prophylaxis (doxyPEP) has been investigated against a number of bacterial STIs. Doxycycline is a widely accessible second-generation tetracycline, known for its tolerability and low risk of severe side effects[6]. A recent meta-analysis of four randomised trials in the United States[7] (DoxyPEP), France [8-9](IPERGAY and DOXYVAC) and Kenya[10] estimated that doxy-PEP is 77% effective against TP infection, and 53% effective against any bacterial STI among GBMSM[11]. Notably, the Kenyan trial, which included only cisgender women, did not find evidence of a treatment effect[10]. However, the authors hypothesised that this could be due to low adherence.

Despite evidence in support of its efficacy, there are concerns about the effect that doxyPEP will have on antimicrobial resistance (AMR) for sexually transmitted or other bacteria. In the DoxyPEP and DOXYVAC trials, doxycycline-resistant *Neisseria gonorrhoeae* isolates increased in doxyPEP users, compared with baseline or standard care groups, respectively[7,9]. Doxycycline resistance in *Staphylococcus aureus* isolated from the oro- and nasopharynx also increased from baseline in the DoxyPEP trial. In addition, there is concern that doxyPEP could select not only for tetracycline resistence, but also resistance to other classes of antibiotic[12-13]. Indeed, a systematic review found some evidence that daily oral tetracycline can result in such resistance, albeit modest and transient[14]. However, currently available trial data are limited by the small number of isolates that have been tested and no trials have explored resistance in *Treponema pallidum*[11].

Setting aside AMR concerns, modelling studies suggest that with an uptake of 10% and an adherence of 80%, doxyPEP could reduce cumulative syphilis incidence by 10% over the next decade in the United States[15]. As such, doxyPEP could be an effective strategy to prevent syphilis diagnoses, requiring treatment of between 9.5 and 31.1 patients for one year to prevent one case, depending on the eligibility criteria established for its prescription[16].

In light of this promising evidence, doxyPEP has already been recommended for GBMSM and transgender women at an elevated risk of acquiring syphilis in the United States and United Kingdom[17-18]. STI PEP refers to the use of antibiotics, including but not limited to doxycycline, for STI post-exposure prophylaxis. There is evidence that prior to being provided at sexual health services in the UK, a significant minority of GBMSM in the UK are already using this as an STI management strategy[19].

Given evidence of early adoption of doxyPEP, it is important to understand the prevalence of and trends in doxyPEP use in GBMSM. While there is evidence that up to 10% of GBMSM taking HIV PrEP or living with HIV report taking antibiotic STI PEP, little more is known about the characteristics of users because analyses are limited by the size of these studiesp[20-24]. This study aims to synthesise data from nine surveys (2019-2024) in England to estimate the prevalence of STI PEP use and clarify the associated demographic and behavioural characteristics. This characterisation will provide a baseline from which future studies can evaluate the impact of doxyPEP provision.

## Methods

### Data Collection

Data from nine different surveys, conducted between 2019 and 2024, was used in this study: the Gay Men’s Sexual Health Survey (GMSHS) 2019 and 2022 (accessed on 29 June 2023), the PrEP Users Survey 2019 and 2020 (accessed on 2 February 2024), the Prevalence of Prophylaxis for STIs (POPS) survey (accessed on 19 February 2024), and four waves of Reducing Inequalities in Sexual Health (RiiSH) survey: RiiSH 2020, RiiSH 2022, RiiSH 2023, and RiiSH 2024, conducted in 2020, 2022, 2023, and 2024, respectively (RiiSH 2020-2023 were accessed on 2 February 2024, while RiiSH 2024 was accessed on 6 January 2025). The researchers did not have access to information that could identify individual participants for any of the surveys. Details regarding the study period, eligibility criteria, sampling method, and data collection method folr each survey are provided in Supplementary Table 1. A description of each variable, including its name in the model is provided in Supplementary Table 2.

### Data Preparation

In order to harmonsise the data from the eight surveys and account for differences in survey questions, composite variables had to be generated. These were *STI diagnosis, chemsex use*, and *history of condomless anal sex*. These were binary variables to indicate whether participants had reported an STI diagnosis, reported havig multiple sexual partners, reported sexualised drug use, or reported having condomless sex, during the survey lookback periods. Notably, survey lookback periods varied from three months to ‘ever’. The *chemsex use* variable also refered specifically to gamma-hydroxybutyrate/gamma-butyrolactone (GHB), methamphetamine, or methadone use.

Observations with missing values for the *age group, gender, ethnicity, and sexual orientation* variables were dropped. In order to reduce data sparcity, the responses *No* and *Not Sure* were combined in the *STI diagnosis* variable. Missing data in the outcome variable (*STI PEP use*) was recoded, assuming that any participants who did not answer this question had not used STI PEP. To address partial missingness across other variables (Table 1), multiple imputation was used to impute the following variables: *education level, HIV status, past HIV PrEP use, STI diagnosis*. Chained logistic regression equations were used to impute missing values (Supplementary Table 3). Ten imputed datasets were generated and a random seed was used to enable reproducibility.

**Table 1.**
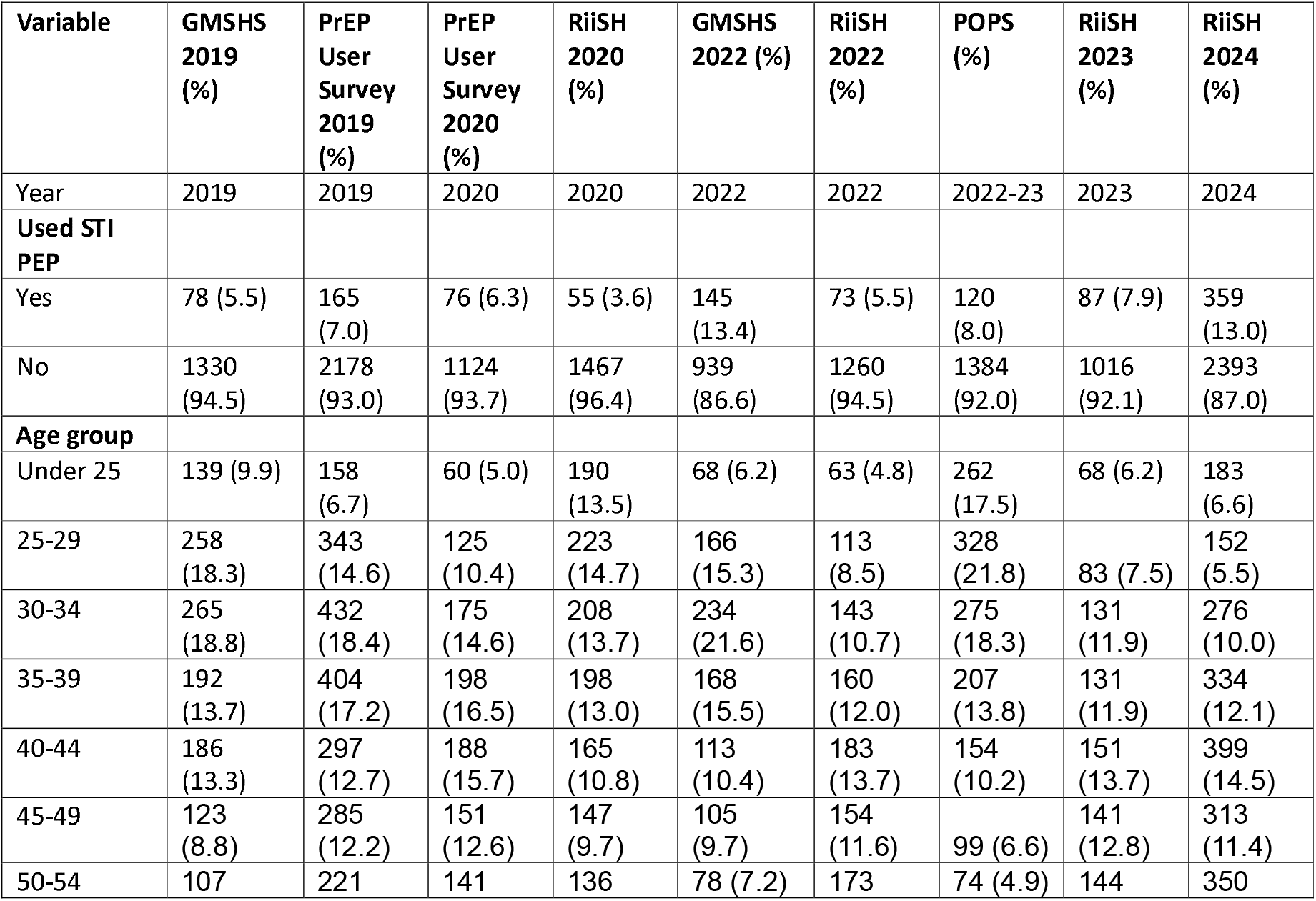
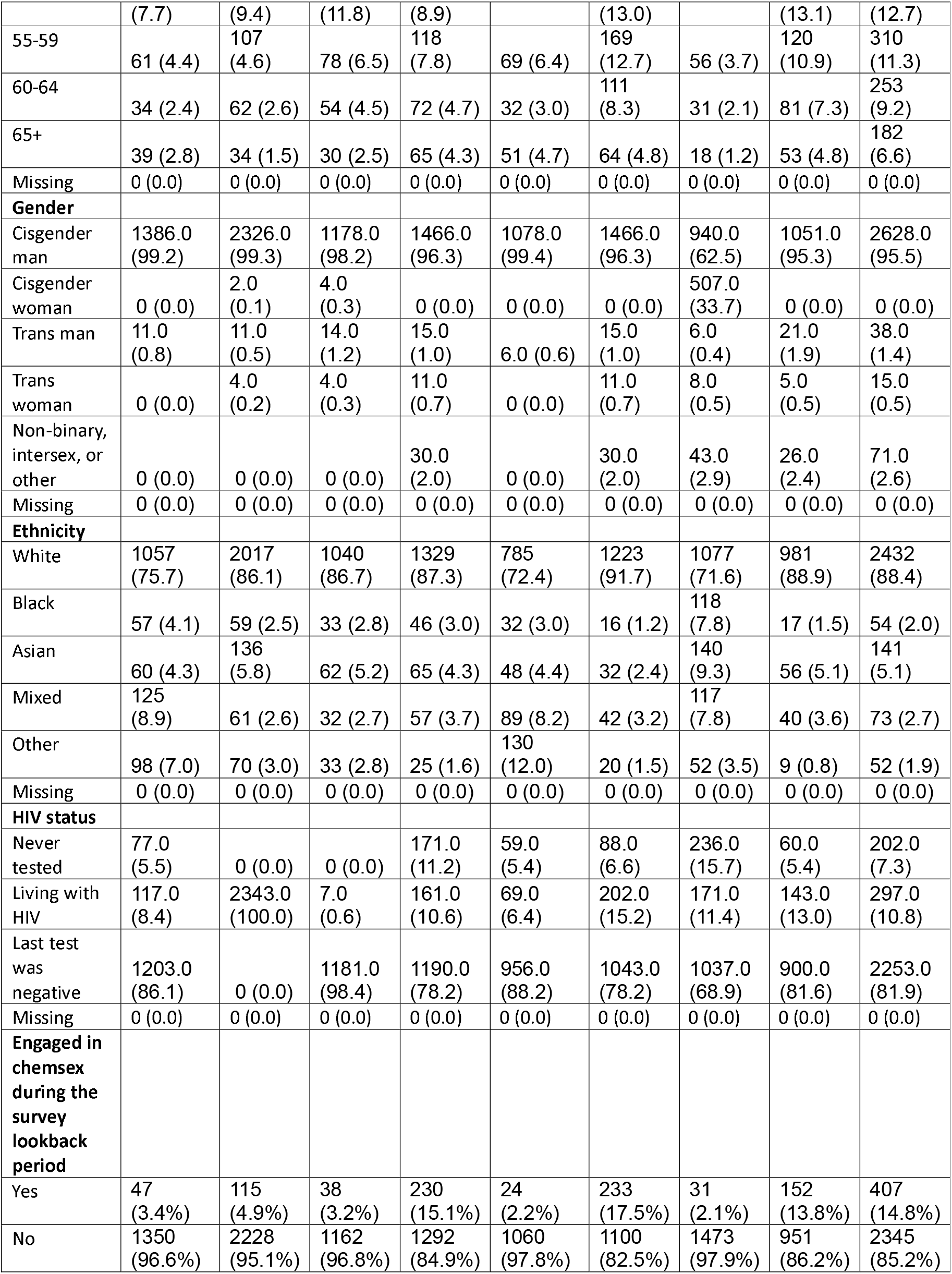
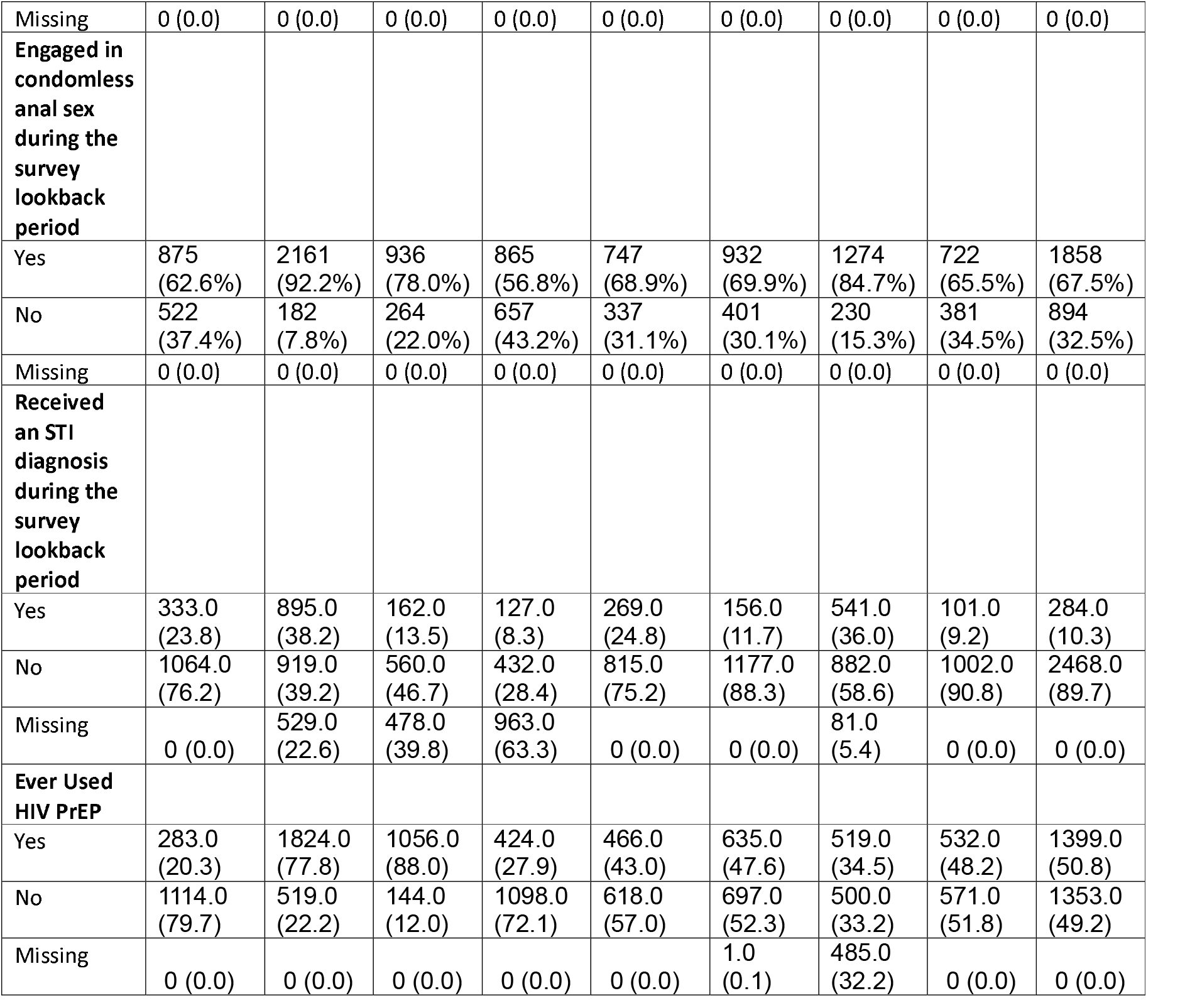
Distribution of demographic and behavioural variables by survey.

### Data Analysis

We calculated the percentage of participants reporting STI PEP use, and the associated 95% confidence intervals, separately for each survey. Factors associated with STI PEP use were modelled using mixed-effects logistic regression. Variables were considered for inclusion if they were considered potentially important or useful predictors for STI PEP use based on existing literature.

All composite variables (*STI diagnosis, chemsex use, history of condomless anal sex*, and *past HIV PrEP use*) were candidates for inclusion as random effects in the full analysis. This was because the exact meaning of the composite variables differed between studies, so we expected heterogeneity in their effects on the outcome (*STI PEP use*). The model thus consists of (1) fixed effects, which are common to all studies, (2) random intercepts, such that baseline prevalence (where all covariates are set to zero) varies between studies, and (3) random covariates, for which the effect of the variable has a mean and variance, and allowed to vary between studies. Each random effects is assumed to have a normal distribution with mean zero and estimated variance, and are assumed to be uncorrelated. To determine which composite variables to include as random effects, we tested for an interaction between each composite variable and the survey on *STI PEP use* in each of the ten imputed datasets.

For each composite variable, two nested models were compared using a likelihood-ratio test. The simpler model included all variables as fixed-effect predictors, including the composite variable of interest. The more complex model included an additional interaction term between the composite variable of interest and survey. If strong evidence of an interaction was found (p<0.05), the composite variable was included as a random effect. Otherwise, the variable was included as a fixed effect.

Data were analysed using STATA 18.0 (College Station, Texas, USA).

## Results

Across the nine surveys, there were a total of 14,142 participants. The distribution of participant characteristics is summarised by survey in Table 1. Prior to multiple imputation, there was partial missingness across several variables. The distribution of this missingness is also summarised in Table 1. The prevalence of STI PEP use varied significantly between surveys, with the highest prevalence observed in the most recent dataset (RiiSH-2024) at 13.05% (95% confidence intervals: 11.85-14.36%) and the lowest observed in RiiSH 2020 at 3.61% (2.78-4.68%). The prevalence of STI PEP use in each survey is reported in Table 1.

Likelihood tests for interactions showed that there was strong statistical evidence of interaction effects (p < 0.05) of *STI diagnosis* and *survey*, as well as *past HIV PrEP use* and *survey* on *STI PEP use* in most of the ten imputed datasets. Despite this interaction, *past HIV PrEP use* was not included as a random effect because it was found to be a perfect predictor of the outcome in PrEP User Survey 2019, so inclusion as a random effect could have resulted in convergence issues. There was no evidence of an interaction effect of *chemsex* or *history of condomless anal sex*, on *STI PEP use* in any of the imputed datasets. The full results of this analysis are reported in Supplementary Table 4.

In light of this, the following variables were included as fixed effects in the final model: survey year, *age group, gender, ethnicity, HIV status, sexual orientation, chemsex use*, and *history of condomless anal sex*. To account for heterogeneity across different surveys, we specified a random intercept for each survey. To account for differences in its definition between surveys, *STI diagnosis* was included as a random effect.

In the full analysis, we found strong statistical evidence of increased STI PEP use among those aged 40-44 (OR = 1.260; 95% confidence interval: 1.000-1.584) and reduced STI PEP use among those under 25 (OR = 0.560; 0.387-0.819) or over 65 (OR = 0.640; 0.407-0.990), compared to those aged 30-34. Transgender women (OR = 3.870; 1.751-8.499) had higher odds of STI PEP use compared to cisgender men. Similarly, Black (OR = 1. 850; 1.336-2.560) and Asian (OR = 1.430; 1.094-1.878) ethnic groups had higher odds compared to those identifying as White. People living with HIV (OR = 3.400; 2.691-4.306) were more likely to use STI PEP than those who last received a negative HIV test.

Additionally, engaging in chemsex (OR = 2.150; 1.804-2.560), condomless anal sex (OR = 1.590; 1.323-1.916), or receiving an STI diagnosis (OR = 1.830; 1.336-2.509) during the lookback period was associated with increased odds of STI PEP use compared to those who had not engaged in these behaviours. There was strong statistical evidence of a substantial increase in the odds of STI PEP use in 2024, compared with 2019 (OR = 4.673; 2.648-8.247). There was no statistical evidence of an increase in any other year, compared with 2019.

The estimated standard deviation for the random slope of STI diagnosis across surveys was 0.398 (95% CI: 0.214 to 0.739), indicating some variability in the association between STI diagnosis and STI-PEP use across different surveys. The random intercept variance across surveys was negligible (SD < 0.001). This suggests that baseline levels of STI PEP use were largely consistent across surveys, once covariates were accounted for. Survey year likely accounted for much of the variance between studies as there were only one or two surveys for each year.

The full results of the mixed-effects logistic regression model are provided in Table 2.

**Table 2.**
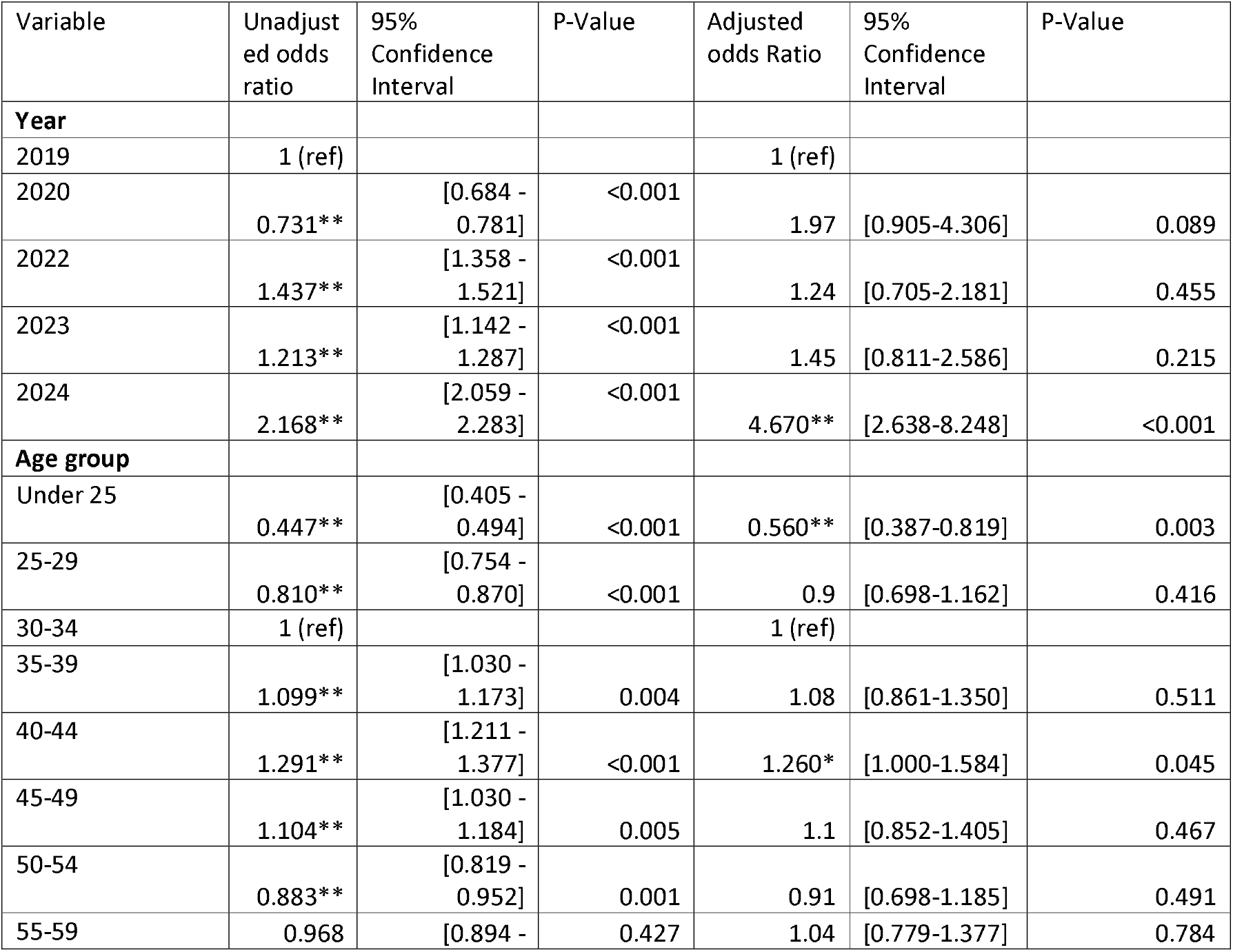
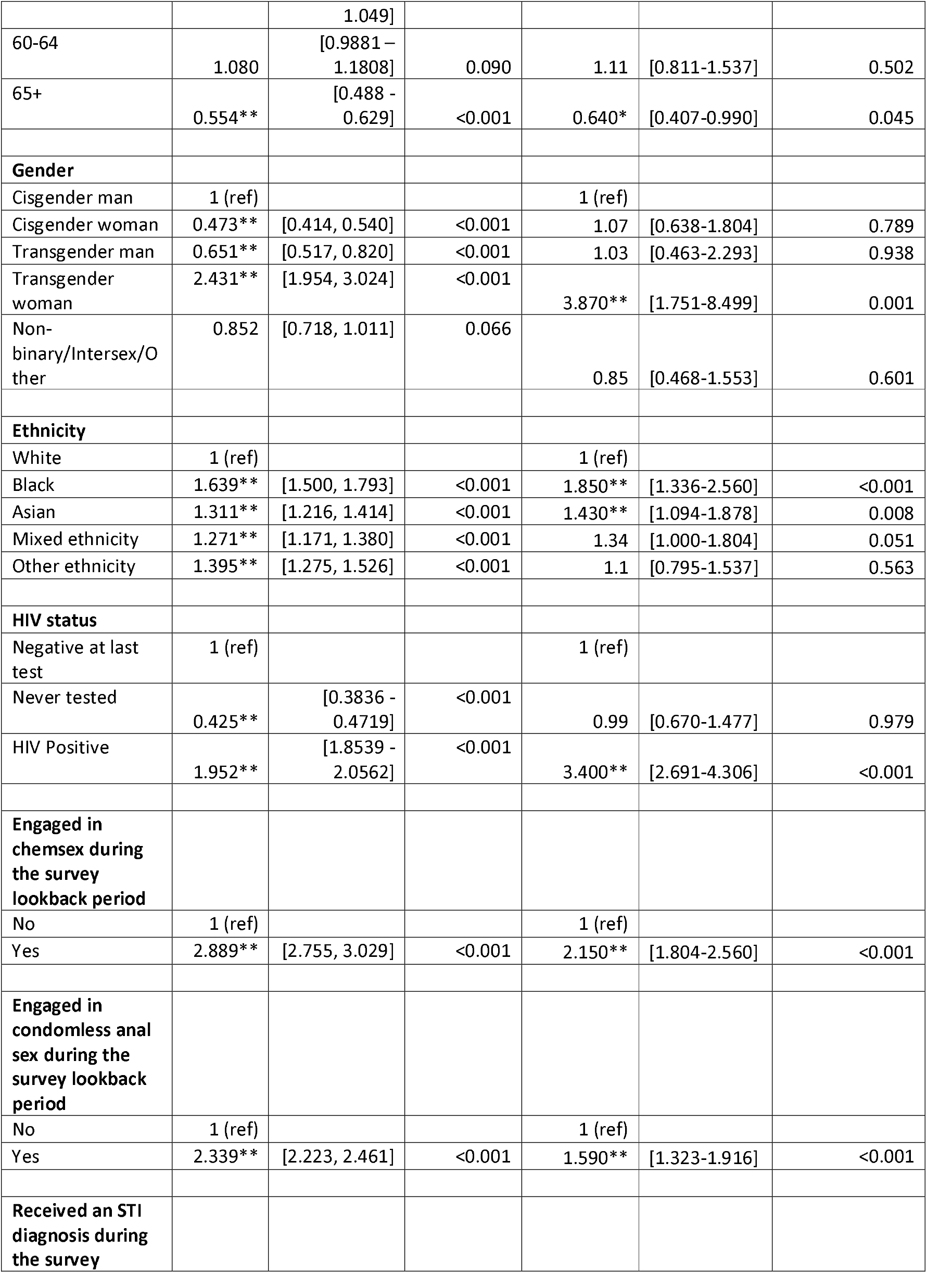
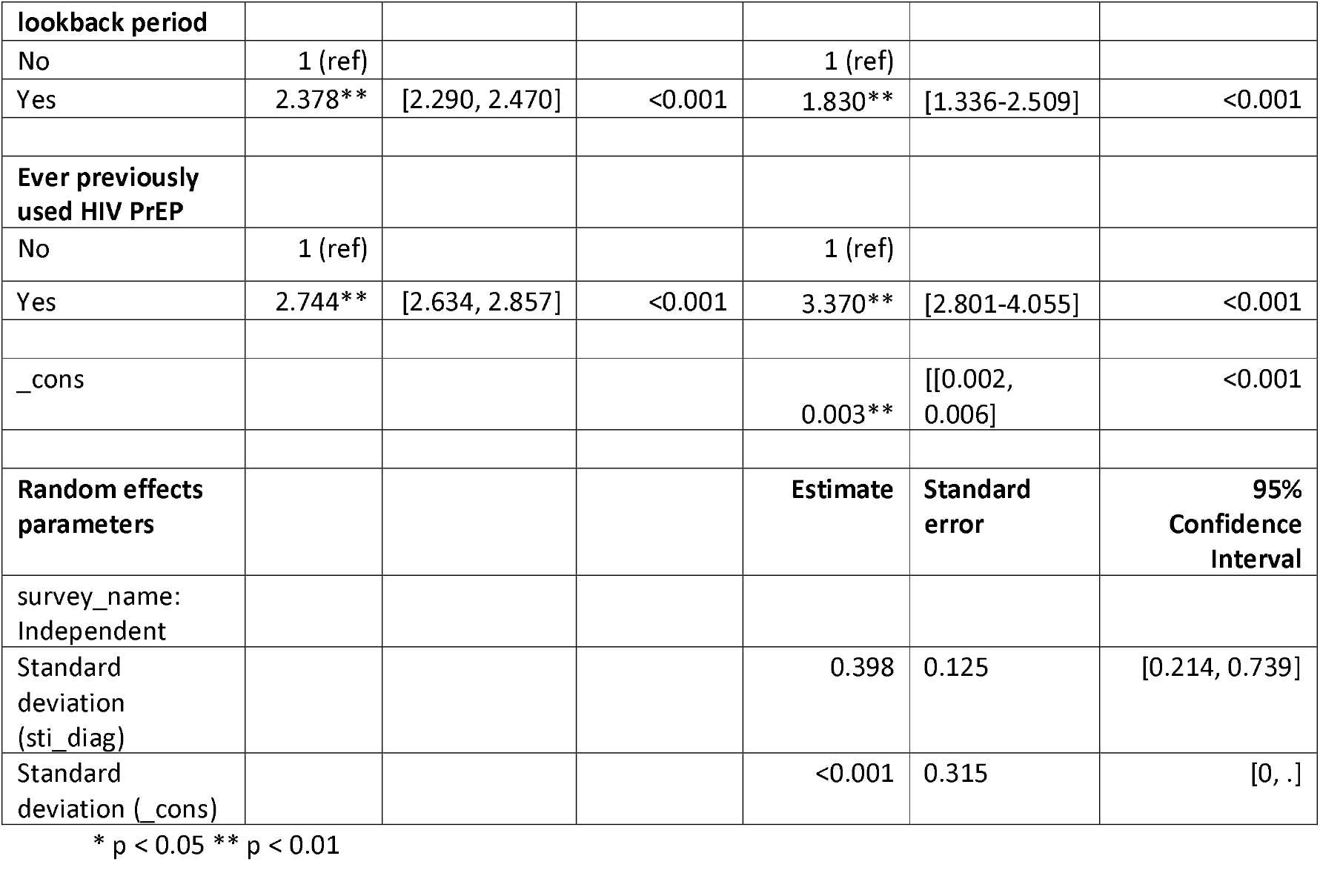
Results from the mixed effects logistic regression model to predict STI PEP use.

## Discussion Main Findings

We synthesised data from nine surveys, conducted in the United Kingdom over five years, to derive an estimate of the prevalence of STI PEP use and build a picture of the factors associated with it.

We observed a wide range of STI PEP use prevalence between surveys(3.6% to 13.4%), which is unsurpising given that the surveys were conducted over a period of five years, with some using different sampling methods and eligibility criteria. Nevertheless, taken together the surveys show that STI PEP is already being used by a significant minority of GBMSM. While some of the STI PEP prevalence estimates provided here have previously been published, this study is unique in providing a sense of the variability of STI PEP prevalence between samples and over time[25].

We also observed associations between year, age, gender, ethnicity, HIV status and behavioural STI risk factors, and STI PEP. Some of these associations were unexpected, while others aligned with previous research.

The sharp increase in the odds of STI PEP use in 2024 may have resulted from increased awareness of STI PEP. Public discourse around STI PEP may have been fueled by the release of national guidlines for doxy-PEP by the US Centres for Disease Control [26].

In terms of ethnicity, the increased odds of STI PEP use among those identifying as Black contradicted our a priori expectations. We would have expected lower uptake among Black individuals, based on the lower HIV PrEP uptake observed in previous research [27-29]. As GBMSM made up the vast majority of our sample, this result likely reflects the association between STI PEP use and being a Black GBMSM, rather than reflecting ethnicity effects in the general population.

In terms of behavioural factors, the association between chemsex, condomless sex, previous STI diagnoses, and the use of STI PEP aligns with findings from previous studies[22, 25, 30]. It also parallels the relationship between behavioural STI risk factors and HIV PrEP use[31]. Together, our findings add to a body of evidence that pharmaceutical STI and HIV prophylaxis is more common among those reporting sexual behaviours which increase their risk of STIs. Importantly, this observation should be caveated with the limited representativeness of our sample.

## Strengths and limitations

This study expands previous research on the factors associated with STI PEP use, by pooling surveys to achieve a greater sample size and statistical power. We have also included the most recent available data in the UK to date. This is a major advantage of our work, as STI PEP is a rapidly expanding practice.

However, our study was limited by the poor representativeness of our sample. Lower response numbers were recorded from young people and people who did not self-describe as white across most surveys. This necessitated combining some non-White ethnic groups in the analysis, limiting the quality findings for minority groups. Surveys also included very few heterosexual people, so the insight we could provide into the prevalence of STI PEP use among them was limited. Given that this group have experienced rising rates of syphilis infection in recent years, this could be viewed as a limitation[2].

Another limitation was the potential for participation bias. The samples in some surveys may have been more engaged in sexual health than the general GBMSM population.

The surveys used in our study are also limited by their cross-sectional methodology. STI PEP use was self-reported so may have been subject to recall bias, for example. Additionally, the absence of information on the chronology of reported behaviours reduces the accuracy with which relationships can be identified, especially in surveys with longer lookback periods.

With regards to our analysis, there were also limitations. We included a random intercept in our model in order to account for differences in the baseline prevalence of STI PEP use across surveys. This was important as surveys were conducted in different years and target populations varied. For example, some surveys were limited to only HIV PrEP users and others were limited to only GBMSM.

Our model also allowed the effect of behavioural risk factors measured using composite variables to vary by survey. This accounted for the fact that the variables had slightly different meanings depending on the survey (for example, the variable describing previous diagnosis with an STI referred to a lookback period of three months in the RiiSH surveys but one year in the GMSHS). We did not, however, allow the effect of other behavioural or demographic factors to vary by survey. This could be problematic given that the surveys were conducted in different years. It is plausible that antibiotic STI PEP use becomes less strongly associated with age as its use becomes more mainstream, for example. A limitation of our model is that it is not sensitive to such varying relationships across all variables.

## Implications for Policy and Practice

Our results indicate that GBMSM at greater risk of STIs—those engaging in chemsex or condomless anal sex, those with a history of STI diagnoses, and those living with HIV—are more likely to use STI PEP. This suggests that the implementation of doxyPEP could achieve high uptake in groups where it would be likely to have more impact, assuming it is used appropriately to maximise benfits (reducing syphilis infection) and minimise potential harms (such as the exacerbation of AMR).

Importantly, our results provide a baseline from which the impact of doxyPEP’s recommendation on it’s uptake can be measured. Such evaluation will be crucial for identifying inequities in doxyPEP implementation.

## Conclusions

By synthesising data from nine different surveys, we derived a comprehensive estimate of who is using STI PEP in the United Kingdom, to date. There is overlap between the characteristics associated with STI PEP use, and those associated with STI risk. However, health disparities almost certainly exist outside of the specific and unrepresentative samples surveyed. To better understand the potential impact of doxyPEP, future surveys should include a more comprehensive sample encompassing all groups at risk of STIs. Nevertheless, these results represent the most granular characterisation of STI PEP users prior to the recommendation of doxyPEP in the UK.

## Data Availability

The data that support the findings of this study contain sensitive personal information and are therefore not publicly available to protect participant privacy. However, some aggregate data may be available upon reasonable request from the UKHSA. Requests can be directed to.

## Ethics statement

Ethical approval of this study was provided by the UKHSA Research Ethics and Governance Group (REGG) - ref: R&D 586. Methods were performed in accordance with guidelines and regulations set by REGG.

