## Supplementary Materials for "An emerging trend in STI Management: Antibiotic STI Post-Exposure Prophylaxis Prevalence and Determinants in Recent Surveys"

**Appendix**

**Supplementary Table 1 – Characteristics of included surveys**

| Survey | Time Period | Eligibility Criteria | Sampling Method | Data Collection |
| --- | --- | --- | --- | --- |
| GMSHS 2019 | 8 June to 17August 2019 | Participants were eligible if they identified as male (including trans men) and self-reported as gay, bisexual, demisexual, pansexual, or having had sex with a man in the last year. | Opportunistic sampling was used. Participants were recruited by fieldworkers at a selection venues (including pubs, clubs, bars and sex-on-premises venues) attended by gay, bisexual and other men who have sex with men. | Surveys were annonymously self-completed by participants, with verbal consent taken by feildworkers. |
| GMSHS 2022 | 4 November 2022 to 4 Feb 2023 | See above. | See above. | See above. |
| PrEP User Survey 2019 | 17 May to 1 July 2019 | Participants were eligible if they were living in the UK and had tried to access or used PrEP since January 2017. | Opportunistic sampling was used. Recruitment occurred via the iWantPrEPNow mailinglist, social media and Grindr. | Online survey. |
| PrEP User Survey 2020 | October to November 2020 | See above. | See above. | See above. |
| POPS | 1 April 2022 to 31 December 2023 | All participants aged 16 or older and able to understand english language were eligible. | Opportunistic sampling was used. Participants were recruited when they attended routine face-to-face or telephone sexual health consultations | Online survey. |
| RiiSH 2020 | 23 Nov 2020 to 12 Dec 2020 | Participants were eligible if they were UK residents, aged 16 or older, identified as male (including trans men) or assigned male at birth (AMAB), and reported having sex in the past year with someone identifying as male or AMAB. | Opportunity sampling was used. Participants were recruited from social networking sites (Facebook, Twitter, Instagram) and dating apps (Grindr and Hornet). | Online survey. |
| RiiSH 2022 | 24 Nov to 19 Dec 2022 | See above. | See above. | See above. |
| RiiSH 2023 | 7 Nov to 6 Dec 2023 | See above. | See above. | See above. |
| RiiSH 2024 | 18 Nov to 10 Dec 2024 | See above. | See above. | See above. |

**Supplementary Table 2 – Description of variables included in the final model**

| **Variable name** | **Description** | **Levels** |
| --- | --- | --- |
| survey_year | Year of survey | 2019 |
|  |  | 2020 |
|  |  | 2021 |
|  |  | 2022 |
|  |  | 2023 |
|  |  | 2024 |
| used_stipep | Used STI PEP | Yes |
|  |  | No |
| age_group | Age group | Under 25 |
|  |  | 25-29 |
|  |  | 30-34 |
|  |  | 35-39 |
|  |  | 40-44 |
|  |  | 45-49 |
|  |  | 50-54 |
|  |  | 55-59 |
|  |  | 60-64 |
|  |  | 65+ |
| gender_full | Combined variable describing gender and sex at birth | Cisgender man |
|  |  | Cisgender woman |
|  |  | Trans man |
|  |  | Trans woman |
|  |  | Non-binary, intersex, or other |
| ethnicity | Ethnicity | White |
|  |  | Black |
|  |  | Asian |
|  |  | Mixed |
|  |  | Other |
| hiv | HIV status | Never tested |
|  |  | Living with HIV |
|  |  | Last test was negative |
| chemsex | Engaged in chemsex during the survey lookback period | Yes |
|  |  | No |
| ever_wcondom | Engaged in condomless anal sex during the survey lookback period | Yes |
|  |  | No |
| sti_diag | Received an STI diagnosis during the survey lookback period | Yes |
|  |  | No |
| used_prep_past | Ever Used HIV PrEP | Yes |
|  |  | No |

**Supplementary Table 3 – Chained conditional models for multiple imputation**

| Imputed (outcome) variable | Model type | Predictor variables included* |
| --- | --- | --- |
| used_prep_past | Multinomial logistic regression (mlogit) | sti_diag, hiv, age_group, gender_full, ethnicity, Ever_wcondom, chemsex |
| sti_diag | Multinomial logistic regression (mlogit) | used_prep_past, hiv, age_group, gender_full, ethnicity, Ever_wcondom, chemsex |
| hiv | Multinomial logistic regression (mlogit) | used_prep_past, sti_diag, age_group, gender_full, ethnicity, Ever_wcondom, chemsex |

* All models were fitted with Stata’s augment option to improve prediction of low-frequency categories.

**Supplementary Table 4 – Likelyhood ratio tests for interactions between survey_name and composite variables**

| Simpler Model | Complex Model | Imputed Dataset | P Value |
| --- | --- | --- | --- |
| logit used_stipep i.age_group i.gender_full i.ethnicity i.hiv chemsex i.survey_name_num | logit used_stipep i.age_group i.gender_full i.ethnicity i.hiv chemsex##i.survey_name_num |  |  |
|  |  | 1 | 0.762 |
|  |  | 2 | 0.749 |
|  |  | 3 | 0.761 |
|  |  | 4 | 0.753 |
|  |  | 5 | 0.755 |
|  |  | 6 | 0.747 |
|  |  | 7 | 0.752 |
|  |  | 8 | 0.762 |
|  |  | 9 | 0.755 |
|  |  | 10 | 0.748 |
| logit used_stipep i.age_group i.gender_full i.ethnicity i.hiv ever_wcondom i.survey_name | logit used_stipep i.age_group i.gender_full i.ethnicity i.hiv ever_wcondom##i.survey_name |  |  |
|  |  | 1 | 0.243 |
|  |  | 2 | 0.249 |
|  |  | 3 | 0.245 |
|  |  | 4 | 0.244 |
|  |  | 5 | 0.246 |
|  |  | 6 | 0.240 |
|  |  | 7 | 0.234 |
|  |  | 8 | 0.241 |
|  |  | 9 | 0.239 |
|  |  | 10 | 0.244 |
| logit used_stipep i.age_group i.gender_full i.ethnicity i.hiv sti_diag i.survey_name | logit used_stipep i.age_group i.gender_full i.ethnicity i.hiv sti_diag##i.survey_name |  |  |
|  |  | 1 | <0.001 |
|  |  | 2 | <0.001 |
|  |  | 3 | <0.001 |
|  |  | 4 | <0.001 |
|  |  | 5 | <0.001 |
|  |  | 6 | <0.001 |
|  |  | 7 | <0.001 |
|  |  | 8 | <0.001 |
|  |  | 9 | <0.001 |
|  |  | 10 | <0.001 |
| logit used_stipep i.age_group i.gender_full i.ethnicity i.hiv used_prep_past i.survey_name | logit used_stipep i.age_group i.gender_full i.ethnicity i.hiv used_prep_past##i.survey_name |  |  |
|  |  | 1 | 0.009 |
|  |  | 2 | 0.059 |
|  |  | 3 | 0.043 |
|  |  | 4 | 0.012 |
|  |  | 5 | 0.016 |
|  |  | 6 | <0.001 |
|  |  | 7 | 0.009 |
|  |  | 8 | 0.009 |
|  |  | 9 | 0.009 |
|  |  | 10 | 0.005 |
